# Evaluating the Reporting Quality of 21,041 Randomized Controlled Trial Articles

**DOI:** 10.1101/2025.03.06.25323528

**Authors:** Apoorva Srinivasan, Jacob Berkowitz, Sophia Kivelson, Nadine Friedrich, Nicholas Tatonetti

## Abstract

Incomplete reporting of a study’s methods and results hinders efforts to evaluate and reproduce research findings in randomized controlled trials (RCTs), leading to potential harm. CONSORT, a set of widely endorsed RCT reporting guidelines, was designed to mitigate such harm by ensuring transparency, reproducibility and safety in RCTs. Evaluating adherence to CONSORT requires a level of language understanding that has previously precluded broad systematic assessment. However, recent advances in large language models (LLMs) now make it possible to evaluate the quality of RCTs at scale. We demonstrate that GPT-4o-mini used out of the box achieves state-of-the-art performance in evaluating RCT quality (F1 score: 0.85; precision: 0.96), with results validated by expert human annotators showing 92.24% agreement across 50 papers. Applying this tool to 21,041 open-access RCTs (1966–2024), we reveal temporal and domain trends: overall CONSORT compliance has improved substantially over time, rising from 27.3% in 1966-1990 to 56.1% in 2010-2024, yet critical methodological components remain severely underreported. Randomization procedures (9.7%), allocation concealment mechanisms (15.25%), and protocol access information (2.22%) were particularly deficient. Substantial variation exists across medical disciplines (35-63% compliance), with urology/nephrology and critical care demonstrating the highest compliance, while pharmacology showed the lowest. Trial characteristics including FDA regulation status, presence of data monitoring committees, reporting of adverse events, and mortality outcomes showed statistically significant but practically negligible differences in compliance rates. Our work provides a scalable AI framework to audit and improve RCT reporting, offering actionable insights for journals, researchers, and policymakers to enhance research integrity and clinical translation.

## 1 Introduction

Randomized controlled trials (RCTs) are the cornerstone of evidence-based medicine. They are used to make regulatory decisions and provide evidence for clinical guidelines and practice. Rigorous design and conduct, combined with complete and accurate reporting, are essential for fully leveraging the strengths of RCTs. However, many randomized controlled trials have methodological flaws and results are often biased Macleod et al. [2014]. Across RCTs, there is a major risk for inflated significance estimates and problems with randomization, allocation concealment, and blinding if certain procedures are not respected Prayle et al. [2012], Ioannidis 2008]. Poor reporting is a persistent issue potentially obscuring biases, complicating replication efforts and ultimately undermining the trustworthiness of biomedical science Simera et al. [2010], Glasziou et al. [2014]. Reporting information about trial methods (e.g., sample size calculation, randomization, and masking) and results (e.g., effect estimates) are necessary to assess validity for use in research, evidence synthesis, and clinical guideline generation. While complete reporting alone is not sufficient to ensure the research is rigorous and reproducible, it is necessary.

The CONSORT Statement, first published in 1996 and updated in 2001 and 2010 Begg et al. [1996], Moher et al. [2001], Schulz et al. [2010], Moher et al. [2010], comprises a 25-item checklist designed to improve completeness and consistency in trial reporting. While CONSORT has been widely endorsed by biomedical journals and extended to various trial designs and interventions Hopewell et al. [2008], Vohra et al. [2015], Boutron et al. [2017], Grant et al. [2018], Junqueira et al. [2023], Kane et al. [2007a], adherence remains inconsistent across the literature.

Traditional assessments of CONSORT compliance have relied on manual review of relatively small publication samples Turner et al. [2012a], limiting large-scale analysis. Automated approaches are needed for comprehensive evaluation, but previous attempts using rule-based algorithms or traditional machine learning performed poorly on this complex task Kilicoglu [2018], Wang et al. [2020]. Large language models (LLMs) offer a promising alternative, with their ability to process complex language structures and capture reporting nuances.

In this paper, we leverage LLMs to evaluate CONSORT reporting in RCT publications at scale. We first validate our approach on the CONSORT-TM dataset Kilicoglu et al. [2021] with expert review, then analyze reporting trends across 21,041 RCTs from 1966-2024.

Our main contributions are:

1. We develop a zero-shot LLM framework achieving state-of-the-art performance on CONSORT compliance assessment, validated through expert review.

2. We conduct large-scale analysis of CONSORT reporting trends across 21,041 RCTs, revealing significant improvements in reporting over time but persistent gaps in critical methodological details. While compliance has increased from 27.3% in 1966-1990 to 56.1% in 2010-2024, substantial variation exists across medical disciplines (35-63% compliance), with no practical differences in the trial characteristics.

## 2 Related Work

Prior NLP approaches to RCT analysis have primarily focused on PICO (Population, Intervention, Comparator, Outcome) classification [Kiritchenko et al., 2010, Wallace et al., 2016, Nye et al., 2018, Brockmeier et al., 2019, Jin and Szolovits, 2020] and risk of bias assessment [Marshall et al., 2016, 2020], with some work on medical abstract classification [Dernoncourt et al., 2017, Jin and Szolovits, 2018, Li et al., 2021]. These efforts, however, did not specifically target comprehensive CONSORT guideline compliance.

Early CONSORT compliance evaluations used rule-based methods [Wang et al., 2020], while recent approaches employed BERT-based models that showed improvements but required extensive labeled data [Kilicoglu et al., 2023]. Initial LLM applications showed promise—a preliminary study using GPT-3.5 reported encouraging results on 30 sports medicine articles [Wrightson et al., 2023], though a subsequent evaluation of GPT-4’s zero-shot capabilities on our target dataset achieved limited performance (F1 score: 0.51) [Jiang et al., 2024]. Our work enhances these efforts through improved prompting techniques that leverage chain-of-thought reasoning.

Research on CONSORT adherence has traditionally relied on manual coding of small article samples across specific disciplines and time periods [Kane et al., 2007b, Turner et al., 2012b, Mills et al., 2005, Ghimire et al., 2012, 2014, Agha et al., 2014, Mbuagbaw et al., 2014, Zhai et al., 2015, Yin et al., 2021]. While some larger studies have examined CONSORT references [Vinkers et al., 2021, Damen et al., 2023], they typically used simple keyword matching rather than detailed compliance assessment. Recent work has begun analyzing methodology sections at scale [Kilicoglu et al., 2023], but comprehensive evaluation across all CONSORT items remains limited.

To our knowledge, we present the first comprehensive assessment of full-text RCT articles using CONSORT guidelines on a dataset of this magnitude, enabling deeper insights into reporting quality trends across the clinical research landscape.

## 3 Methods

### 3.1 Dataset

For model evaluation, we used a previously curated dataset called the CONSORT-TM corpus [Kilicoglu et al., 2021] for this study. It consists of 50 RCT publications annotated at the sentence level with 37 CONSORT checklist items. In this work, they excluded the checklist item Background (2a) because it is too broad a category that virtually all papers report and checking its reporting was deemed unnecessary. Six annotators independently annotated 30 articles in pairs, and the calculated Krippendorff’s *α* to measure inter-annotator agreement at the sentence level was 0.57. The dataset is available on GitHub https://github.com/ScienceNLP-Lab/RCT-Transparency1.

For large-scale analysis, we identified 53,137 open-access human RCTs from PubMed (1966-2024) and successfully obtained 21,041 full-text PDFs. Articles spanned four time periods: 1966-1990 (*n* = 2, 771), 1990-2000 (*n* = 1, 969), 2000-2010 (*n* = 3, 765), and 2010-2024 (*n* = 10, 447). PDFs were converted to XML using PyMuPDF and enriched with metadata via Semantic Scholar. For a subset (*n* = 1, 790), we extracted NCT numbers and obtained trial characteristics from ClinicalTrials.gov, including phase, funding source, FDA status, data monitoring committees, and safety outcomes. The complete dataset of 21,041 RCT assessments has been made publicly available for research purposes here: https://github.com/ScienceNLP-Lab/RCT-Transparency

### 3.2 Model Architecture

To extract methods-related CONSORT checklist items from RCT reports for this study, we consider OpenAI’s proprietary models GPT-4, GPT-4o and GPT-4o-mini. We run the GPT models via a secure Azure PHI-compliant instance.

We begin by evaluating the zero-shot capabilities of our LLMs. We choose the zero-shot setting as it offers the simplest path towards real-world deployment – it requires minimal engineering, no data labeling, and it can instantly be adapted to any RCT.

Each criterion was assessed independently for every article. For each criterion, the entire article content was fed into the model, and the assessment was conducted one criterion at a time. As a result, the model must be prompted with the same article *C* times, where *C* is the number of criteria.

The output that we expect from the model is a JSON string containing four elements:

1. Criterion: The specific criterion being assessed
2. Rationale: Step-by-step reasoning as to why the patient does or does not meet the criterion
3. Decision: Output “MET” if the patient meets the criterion, or it can be inferred that they meet the criterion with common sense. Output “NOT MET” if the patient does not, or it is impossible to assess given the provided information
4. Confidence: “Low”, “Medium”, or “High” confidence in prediction

The rationale provided by the LLM was particularly important for model explainability, as it offered a detailed explanation of the decision-making process for each criterion. Additionally, such exposed chain-of-thought approaches are known to significantly increase LLM accuracy on a range of language understanding tasks. Self-reported confidence levels were used to assess the reliability of the model’s output, helping us determine when manual intervention might be required for borderline or ambiguous cases.

### 3.3 Model Evaluation and Validation

We evaluated the ability of our LLMs to determine whether an RCT article met or did not meet a set of inclusion criteria for each of the 25 CONSORT items. The models were assessed based on their performance across standard binary classification metrics, including precision, recall, and both macro and micro F1 scores. To address potential hallucination risks, we analyzed performance stratified by model-reported confidence levels and conducted human validation using four experts (one clinician, three data scientists) who manually evaluated outputs from 50 randomly selected articles as Correct/Partially Correct/Incorrect.

### 3.4 Model Application

We applied our most efficient and best performing model to 21,041 open access NCBI articles published between 1966-2024 to evaluate the quality of RCT reporting over time.

For each article, the model was tasked with independently assessing each criterion. This required re-prompting the model for every criterion, ensuring that each article was evaluated multiple times—once for each criterion.

### 3.5 Large-scale Analysis

We applied our best-performing model to assess CONSORT compliance across 21,041 RCTs. We conducted temporal trend analysis across four publication periods, disciplinary analysis by mapping journals to Scimago categories, and relationships with trial characteristics from ClinicalTrials.gov data.

We analyzed reporting patterns across CONSORT section categories (Title & Abstract, Introduction, Methods, Results, Discussion, Other Information) and assessed reporting quality variation by trial phase, funding source, FDA status, geographic location, safety monitoring, and adverse event reporting. We also examined correlations between reporting quality and citation impact using both total and influential citation metrics from Semantic Scholar.

Statistical significance was evaluated using chi-square tests for categorical comparisons and Pearson correlation for continuous relationships, with effect sizes calculated using Cramer’s V. All analyses were performed using Python 3.8 with scipy.stats and statsmodels packages.

## 4 Results

### 4.1 Zero-shot Criteria Assessment

All GPT models with our prompting scheme substantially outperformed the previous state-of-the-art on CONSORT-TM by over 40% [Damen et al., 2023]. GPT-4 achieved the highest F1 score (0.89) and strong precision (0.93). GPT-4o maintained strong precision (0.94) with a slightly lower F1 score (0.85), while GPT-4o-mini demonstrated the highest precision (0.96) with equivalent F1 performance (0.85). Given its balance of accuracy and computational efficiency, we selected GPT-4o-mini as our deployment model.

**Table 1:** Zero shot results on CONSORT-TM dataset

| Model | Accuracy | Precision | Recall | F1 Score | Micro F1 Score |
| --- | --- | --- | --- | --- | --- |
| GPT-4-turbo | 0.84 | 0.93 | 0.85 | 0.89 | 0.84 |
| GPT-4o | 0.80 | 0.94 | 0.79 | 0.85 | 0.80 |
| GPT-4o-mini | 0.81 | 0.96 | 0.77 | 0.85 | 0.81 |
| Lan Jiang et al. | - | 0.48 | 0.54 | 0.51 | 0.49 |

### 4.2 Model Validation

Model performance strongly correlated with self-assessed confidence levels. High-confidence predictions (majority of assessments) achieved excellent performance (F1: 0.95, precision: 0.97), while medium-confidence predictions showed markedly lower reliability (F1: 0.31, precision: 0.85). No predictions were made with low confidence, suggesting effective calibration of the confidence scoring mechanism. Based on this clear performance stratification, we restricted all subsequent analyses to high-confidence predictions only. This approach enabled us to maximize both precision and coverage while systematically excluding potentially unreliable judgments where the model detected ambiguity or insufficient information in the text. The resulting filtered dataset maintained robust coverage (*>*90% of articles) while significantly enhancing assessment reliability.

**Table 2:** Model performance metrics stratified by confidence levels

| Confidence | Accuracy | Precision | Recall | F1 Score | Micro F1 Score |
| --- | --- | --- | --- | --- | --- |
| High | 0.92 | 0.97 | 0.92 | 0.95 | 0.92 |
| Medium | 0.41 | 0.85 | 0.19 | 0.31 | 0.42 |

Human expert evaluation of GPT-4o-mini rationales for 50 articles showed 83.42% correct, 8.82% partially correct, and 7.76% incorrect assessments. Error analysis revealed two main limitations: (1) misclassification of absent events (e.g., no protocol changes) as unreported items, and (2) incorrect inference of study locations from author affiliations.

Based on human expert validation, we identified four CONSORT items that our model had difficulty accurately assessing: interim analyses and stopping guidelines (Item 7b), changes to methods after trial commencement (Item 3b), changes to trial outcomes (Item 6b), and reasons for trial termination (Item 14b). These items share a common characteristic—they report events that may not occur in every trial (e.g., not all trials have protocol changes or early termination). Our model often misinterpreted the absence of these events as non-reporting rather than recognizing them as not applicable. Additionally, these items were infrequently reported across the dataset (reported in *<*5% of articles), limiting their impact on overall compliance scores. Therefore, we excluded these four items from our final analysis to ensure the reliability of our findings.

### 4.3 Large-scale RCT Reporting Quality Analysis

#### 4.3.1 Overall CONSORT Item Reporting

The most frequently reported CONSORT items were scientific background/rationale (95.88%) and specific objectives/hypotheses (89.21%). However, critical methodological details were often missing. Only 1.6% discussed external validity, 9.7% reported randomization sequence generation, 15.25% described allocation mechanisms, and 2.22% provided protocol access information (Fig. 2A).

**Figure 1.**
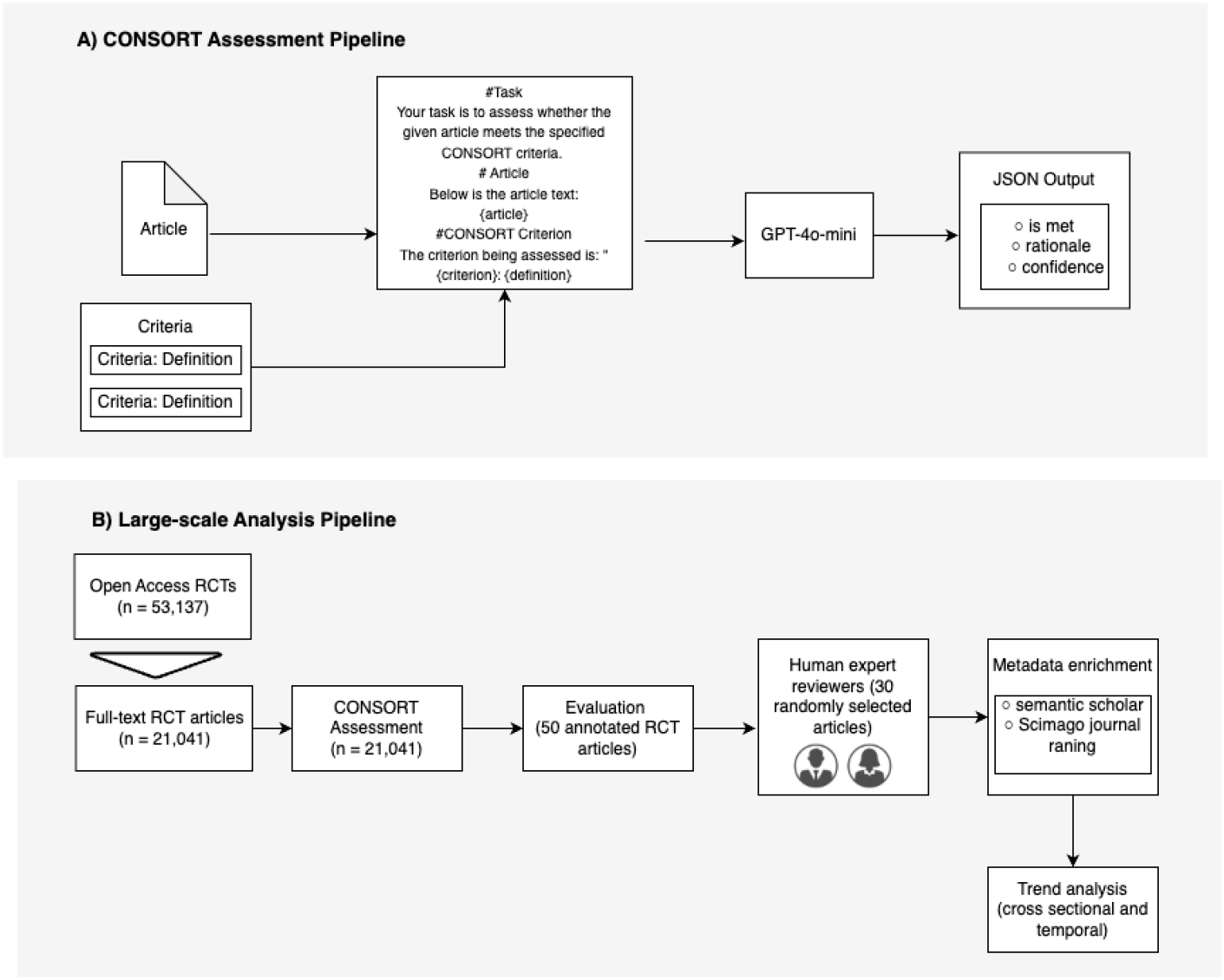
System architecture for automated CONSORT adherence assessment. (a) Zero-shot assessment pipeline: Research articles are evaluated against CONSORT criteria using GPT-4o-mini, producing structured JSON outputs with decisions, rationales, and confidence scores. (b) Large-scale analysis framework: The validated system processes 21,041 RCTs, with results verified by expert review and enriched with metadata for trend analysis.

**Figure 2.**
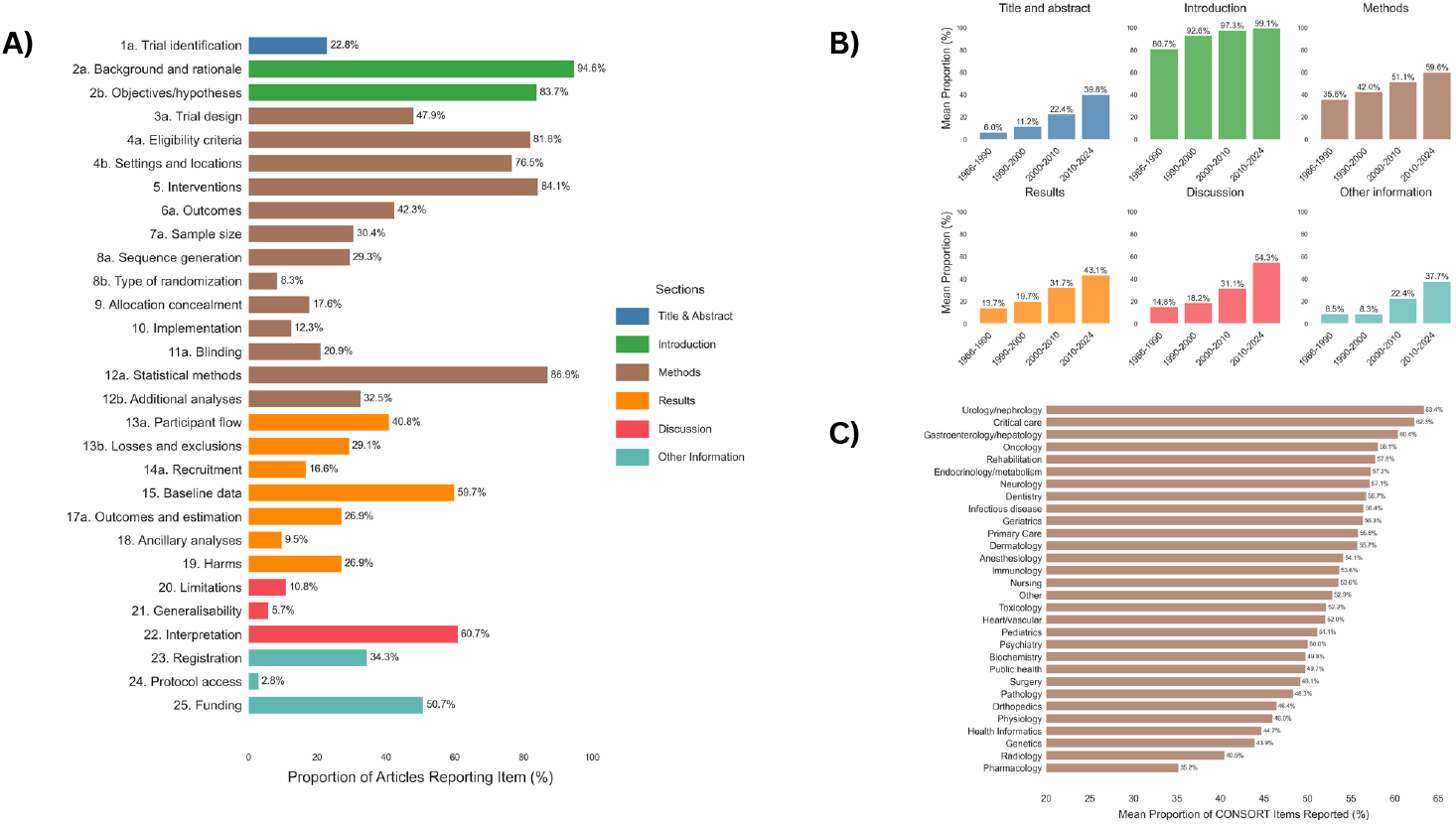
CONSORT Reporting Analysis Across Scientific Literature. (A) CONSORT items reporting rates in randomized controlled trials (*n* = 21, 041). Bars represent the proportion of articles that adequately report each CONSORT checklist item, color-coded by manuscript section (Title & Abstract, Introduction, Methods, Results, Discussion, Other Information); (B) Temporal trends in CONSORT reporting by article section (1966-2024). Each panel shows the evolution of reporting compliance across four time periods, demonstrating substantial improvement across all sections, with particularly notable gains in Results, Discussion, and Other Information (which includes details such as trial registration and funding sources) sections; (C) CONSORT reporting rates by medical discipline. The highest reporting quality was observed in specialized fields such as urology/nephrology and critical care (*>*62%), while pharmacology showed the lowest adherence (35.2%). Data suggest marked variability in CONSORT guideline implementation across medical specialties.

#### 4.3.2 Temporal Trends in Reporting

We observed substantial improvement in CONSORT reporting rates over time (Fig. 2B). The mean compliance rate increased from 27.3% (95% CI: 27.0-27.6%) in 1966-1990 to 33.9% (95% CI: 33.5-34.3%) in 1990-2000, representing a 24.3% relative increase (*p <* 0.0001). This upward trend continued with reporting rates rising to 45.0% (95% CI: 44.7-45.3%) in 2000-2010, a 32.7% increase from the previous decade (*p <* 0.0001). The most recent period (2010-2024) showed further improvement to 56.1% (95% CI: 56.0-56.3%), representing a 24.7% increase (*p <* 0.0001). Despite these significant improvements across each time interval, the average proportion of CONSORT items reported remained below 60% even in the most recent period, indicating persistent reporting gaps despite widespread guideline adoption.

#### 4.3.3 Variations by Field and Discipline

The reporting of CONSORT items varied significantly across medical disciplines (Fig. 2C). Urology/nephrology had the highest proportion of items met over the full-time interval measured, with 63.35% of criteria reported, followed by critical care at 62.27% and Gastroenterology/hepatology at 60.28%. On the lower end of the spectrum, Pharmacology had the lowest reporting rates, with only 35.16% of items met, followed by Radiology (40.46%).

#### 4.3.4 Trial Characteristics and Oversight

##### Trial Phase and Funding

Early phase 1 (57.9%) and phase 1 (59%) trials showed the lowest reporting rates, while phase 2 (66.56%) trials demonstrated the highest compliance followed by phase 4 (64.87%) (Fig. 3A). Regarding funding sources, while individual investigators (72.3%) showed the highest compliance, their small sample size limits interpretation. Among major funders, federal agencies (66.1%) and industry-sponsored trials (63.8%) showed similar compliance rates with no significant difference between them. Network-sponsored trials and other government agencies showed moderate compliance rates at 64.8% and 62.5% respectively (Fig. 3C).

**Figure 3.**
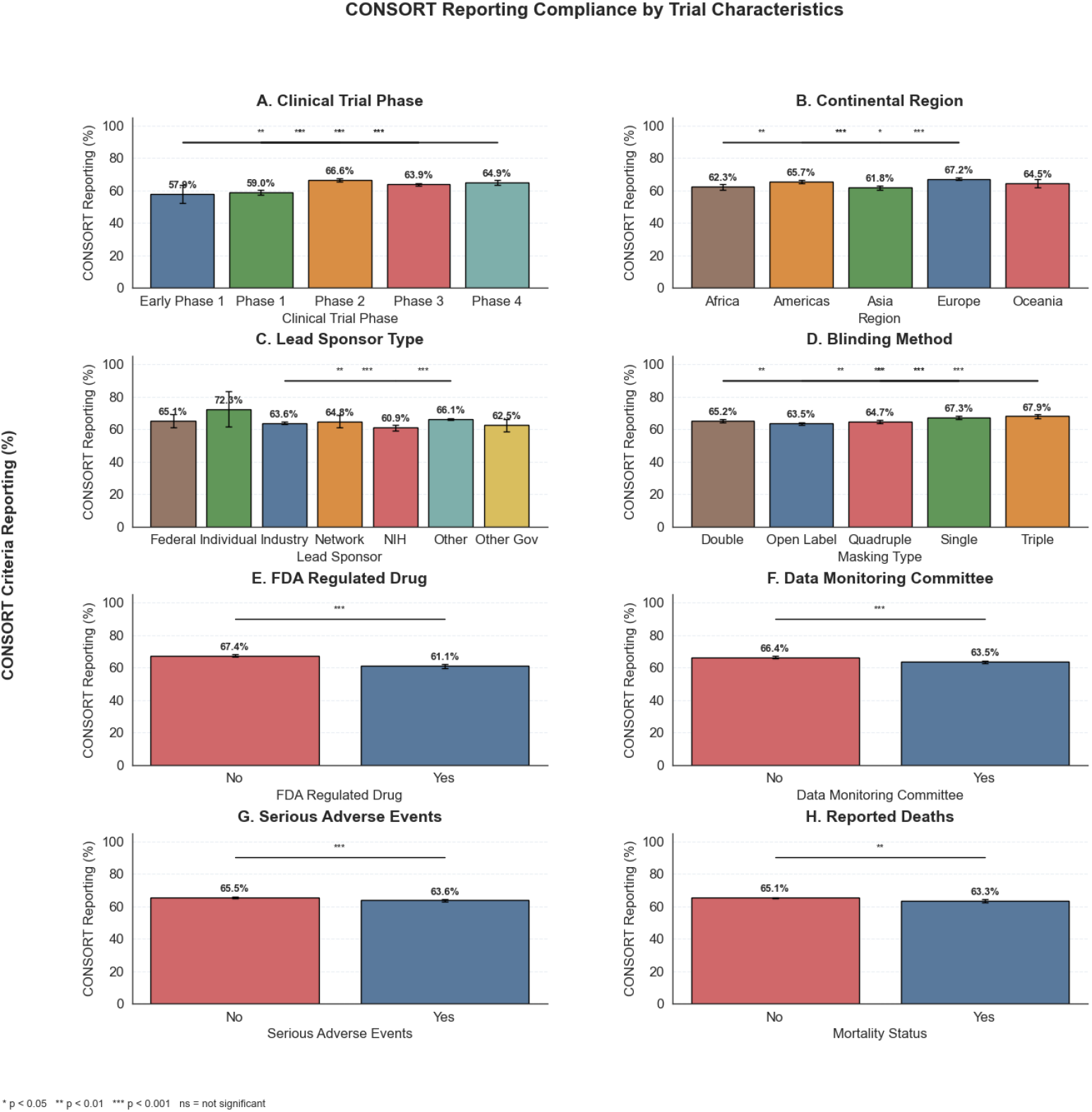
CONSORT Reporting Compliance Stratified by Trial Characteristics. Reporting adherence to CONSORT guidelines across diverse trial attributes: (A) Clinical trial phase, with Phase 2 trials showing highest compliance (66.6%); (B) Continental region, with Europe demonstrating strongest reporting (67.2%); (C) Lead sponsor type, with individual investigators showing highest compliance (72.5%) among categories; (D) Blinding method, showing higher compliance in triple-blind (67.9%) and single-blind (67.3%) trials; (E) FDA regulated status, with non-FDA-regulated trials showing higher compliance (67.4% vs 61.1%); (F) Data monitoring committee presence, showing higher compliance in trials without DMCs (66.4% vs 63.5%); (G) Serious adverse events, showing higher compliance in trials without reported SAEs (65.5% vs 63.6%); and (H) Mortality outcomes, showing higher compliance in trials without reported deaths (65.1% vs 63.3%). Asterisks indicate statistical significance: \**p <* 0.05, \*\**p <* 0.01, \*\*\**p <* 0.001; ns = not significant.

##### Regulatory Oversight and Safety Monitoring

FDA-regulated trials showed lower compliance (61.1%) compared to non-FDA-regulated trials (67.4%, *p <* 0.001, Cramer’s V=0.06) (Fig. 3E). Trials with Data Monitoring Committees showed lower compliance (63.5%) than those without (66.4%, *p <* 0.001, Cramer’s V=0.03) (Fig. 3F). Similarly, trials reporting serious adverse events (63.6% vs 65.5%, *p <* 0.001, Cramer’s V=0.02) or deaths (63.3% vs 65.1%, *p <* 0.01, Cramer’s V=0.01) showed slightly lower compliance rates (Fig. 3G-H). This pattern consistently suggests that characteristics typically associated with increased trial oversight and safety monitoring were paradoxically associated with lower CONSORT reporting compliance, though the very small effect sizes (all Cramer’s V *<* 0.10) indicate these differences have limited practical significance despite their statistical significance.

#### 4.3.5 Citation Impact

CONSORT reporting completeness showed weak but statistically significant correlations with both total citation counts (*r* = 0.062, *p <* 0.001) and influential citations (*r* = 0.101, *p <* 0.001). While regression analysis confirmed these relationships (*p <* 0.001), the proportion of variance explained was modest (*R*^2^ = 0.004 for total citations, *R*^2^ = 0.010 for influential citations), suggesting limited practical significance.

## 5 Discussion and Future Work

Our study demonstrates that large language models can achieve state-of-the-art performance in evaluating CONSORT compliance through zero-shot prompting, without requiring extensive fine-tuning. The GPT-4o-mini model achieved excellent performance (F1: 0.85; precision: 0.96) with high agreement with human experts (92.24%), validating the potential of LLMs for automated assessment of clinical trial reporting quality at scale.

Analyzing over 21,000 RCTs spanning nearly six decades revealed several important patterns in CONSORT compliance. While reporting quality has improved substantially since 2010—coinciding with updated CONSORT guidelines and increasing journal adoption—significant gaps persist in critical methodological details. The improvement from 27.3% compliance in pre-1990 publications to 56.1% in the 2010-2024 period reflects growing awareness of reporting standards yet remains far from optimal. Most concerning was the systematic underreporting of elements crucial for trial reproducibility and methodological integrity, including randomization procedures (9.7%), allocation concealment mechanisms (15.25%), and protocol access information (2.22%). These gaps limit the ability of researchers, clinicians, and systematic reviewers to fully evaluate the validity of reported findings.

The substantial variation in compliance across medical disciplines (ranging from 35% to 63%) likely reflects differences in research culture, methodological traditions, and journal policies across specialties. Fields with strong methodological traditions and established research networks, such as urology/nephrology and critical care, demonstrated exemplary reporting practices. Conversely, pharmacology and radiology showed concerning gaps, suggesting that certain disciplines may benefit from targeted educational interventions. These disciplinary differences persist even after accounting for publication year, indicating that factors beyond temporal trends influence reporting quality.

Our analysis of trial characteristics revealed that later-phase trials showed better compliance than early-phase studies, suggesting that established protocols and increased experience with a particular intervention may improve reporting practices. However, we observed several counterintuitive patterns in relation to oversight mechanisms. FDA-regulated trials, trials with data monitoring committees, and trials reporting serious adverse events or deaths showed slightly lower CONSORT compliance compared to those without these characteristics. While these differences were statistically significant, the very small effect sizes indicate limited practical significance. These patterns may reflect complex reporting environments where regulatory submissions and safety monitoring exist in parallel to, rather than integrated with, academic publication practices.

Our findings have important implications for multiple stakeholders in the clinical research ecosystem. For journal editors and publishers, they highlight the need for more rigorous enforcement of CONSORT guidelines during peer review and editing processes, particularly for consistently underreported items. Implementing automated pre-submission checks using approaches like ours could provide authors with immediate feedback before peer review, potentially improving reporting quality.

For researchers, our results identify specific reporting elements that require greater attention. The persistent underreporting of randomization procedures, allocation concealment, and protocol information suggests that authors may benefit from more explicit guidance on these elements. Educational interventions targeting these commonly missed items could improve compliance, particularly in disciplines with lower overall adherence.

For funding agencies and regulatory bodies, our findings suggest that regulatory oversight alone does not ensure comprehensive reporting. Integration of academic reporting standards into regulatory requirements could help bridge this gap. Funders might consider requiring CONSORT compliance as part of their grant conditions, particularly for late-phase trials.

For AI development, our successful implementation of a zero-shot LLM approach demonstrates the potential for automated tools to assess research quality at scale. This methodology could be extended to other reporting guidelines (e.g., PRISMA for systematic reviews, STROBE for observational studies) and integrated into manuscript submission systems to provide real-time feedback to authors.

Although our study has numerous strengths, several limitations warrant consideration. First, while we mitigated LLM hallucination risks through confidence-based filtering and expert validation, further refinement of uncertainty quantification methods is needed. Second, our analysis was limited to open-access articles, which may not be representative of all published RCTs. Third, our assessment focused on the presence of reporting elements rather than their quality or accuracy. Finally, evolving reporting standards over time may affect the interpretation of temporal trends, although our methodology attempted to account for this by evaluating articles against consistent criteria.

Future work should focus on several key areas. First, enhancing LLM assessment tools through improved uncertainty quantification could further increase reliability. Second, investigating causal factors underlying reporting disparities across trial phases, funding sources, and disciplines could inform targeted interventions. Third, expanding analysis to emerging guidelines, such as the 2022 CONSORT-Harms update, would provide insights into evolving reporting practices. Fourth, developing and testing interventions based on automated feedback could determine whether real-time assessment improves reporting quality.

Additionally, integration of our approach with journal submission systems could create a practical tool for authors, reviewers, and editors. Such systems could provide immediate feedback on CONSORT compliance, potentially improving reporting quality before peer review. Extending this methodology to other reporting guidelines would further enhance research transparency across study designs.

## Data Availability

All data produced are available online at https://huggingface.co/datasets/apoorvasrinivasan/CONSORT-21K

## Supplementary Appendix

### Table of Contents

1. CONSORT Items
2. Prompt Structure for LLM Assessment
3. Item-level Performance Results
4. Methodology for RCT Identification and Metadata Collection
5. Figures and Tables

### List of Figures and Tables

- Figure S1. Zero-Shot Prompting Framework for CONSORT Compliance Assessment
- Figure S2. Relationship Between CONSORT Compliance and Citation Impact
- Table S1. Comprehensive List of CONSORT Checklist Items by Section
- Table S2. Detailed Model Performance Metrics for Individual CONSORT Items
- Table S3. CONSORT Reporting Compliance by Medical Discipline
- Table S4. Overall Reporting Rates for Individual CONSORT Items
- Figure S3. Validation of LLM Assessment Against Human Expert Evaluation
- Figure S4. Geographic Distribution of CONSORT Reporting Compliance
- Figure S5. CONSORT Reporting Quality by Clinical Trial Phase

### 1. CONSORT Items

**Table S1. CONSORT items definitions, corresponding item numbers, and the sections to which they typically belong**. This table presents the complete list of 33 CONSORT items used in our assessment, organized by their standard item numbers and manuscript sections. These items represent the essential elements that should be reported in randomized controlled trials to ensure transparency and reproducibility.

**Table S1.** Complete list of CONSORT items used for RCT quality assessment

| Criteria | Criteria Section | Criteria Definition |
| --- | --- | --- |
| 1a | Title and abstract | Identification as a randomized trial in the title |
| 2a | Introduction | Scientific background and explanation of rationale |
| 2b | Introduction | Specific objectives or hypotheses |
| 3a | Methods | Description of trial design (such as parallel, factorial) including allocation ratio |
| 3b | Methods | Important changes to methods after trial commencement (such as eligibility criteria), with reasons |
| 4a | Methods | Eligibility criteria for participants |
| 4b | Methods | Settings and locations where the data were collected |
| 5 | Methods | The interventions for each group with sufficient details to allow replication, including how and when they were actually administered |
| 6a | Methods | Completely defined pre-specified primary and secondary outcome measures, including how and when they were assessed |
| 6b | Methods | Any changes to trial outcomes after the trial commenced, with reasons |
| 7a | Methods | How sample size was determined |
| 7b | Methods | When applicable, explanation of any interim analyses and stopping guidelines |
| 8a | Methods | Method used to generate the random allocation sequence |
| 8b | Methods | Type of randomisation; details of any restriction (such as blocking and block size) |
| 9 | Methods | Mechanism used to implement the random allocation sequence (such as sequentially numbered containers), describing any steps taken to conceal the sequence until interventions were assigned |
| 10 | Methods | Who generated the random allocation sequence, who enrolled participants, and who assigned participants to interventions |
| 11a | Methods | If done, who was blinded after assignment to interventions (for example, participants, care providers, those assessing outcomes) and how |
| 12a | Methods | Statistical methods used to compare groups for primary and secondary outcomes |
| 12b | Methods | Methods for additional analyses, such as subgroup analyses and adjusted analyses |
| 13a | Results | For each group, the numbers of participants who were randomly assigned, received intended treatment, and were analysed for the primary outcome |
| 13b | Results | For each group, losses and exclusions after randomisation, together with reasons |
| 14a | Results | Dates defining the periods of recruitment and follow-up |
| 14b | Results | Why the trial ended or was stopped |
| 15 | Results | A table showing baseline demographic and clinical characteristics for each group |
| 17a | Results | For each primary and secondary outcome, results for each group, and the estimated effect size and its precision (such as 95% confidence interval) |
| 18 | Results | Results of any other analyses performed, including subgroup analyses and adjusted analyses, distinguishing pre-specified from exploratory |
| 19 | Results | All important harms or unintended effects in each group |
| 20 | Results | Trial limitations, addressing sources of potential bias, imprecision, and, if relevant, multiplicity of analyses |
| 21 | Discussion | Generalisability (external validity, applicability) of the trial findings |
| 22 | Discussion | Interpretation consistent with results, balancing benefits and harms, and considering other relevant evidence |
| 23 | Discussion | Registration number and name of trial registry |
| 24 | Other information | Where the full trial protocol can be accessed, if available |
| 25 | Other information | Sources of funding and other support (such as supply of drugs), role of funders |

### 2. Prompt Structure for LLM Assessment

**Figure S1. Zero-Shot Prompting Framework for CONSORT Compliance Assessment** The prompt design used for our zero-shot LLM assessment framework is detailed below. This specific prompt structure was critical to achieving state-of-the-art performance in CONSORT compliance evaluation without requiring any fine-tuning. The prompt combines task definition, article content, CONSORT criterion specifications, and structured output formatting to guide the model in performing consistent, high-quality assessments.

### 3. Item-level Performance Results

**Table S2. Detailed Model Performance Metrics for Individual CONSORT Items** This table presents the comprehensive performance evaluation of our zero-shot LLM framework on the CONSORT-TM dataset, broken down by individual CONSORT items. The metrics include precision, recall, F1 score, and accuracy for each of the 33 CONSORT checklist items, demonstrating the model’s strengths and limitations across different reporting elements. These results reveal that certain methodological items (like randomization details) were more challenging to assess than descriptive elements (like background and objectives).

### 4. Methodology for RCT Identification and Metadata Collection

**Comprehensive Pipeline for RCT Data Acquisition and Processing** Our systematic approach to identifying, collecting, and processing RCT publications involved a multi-step pipeline designed to ensure comprehensive coverage and high-quality data extraction:

**Figure S1:**
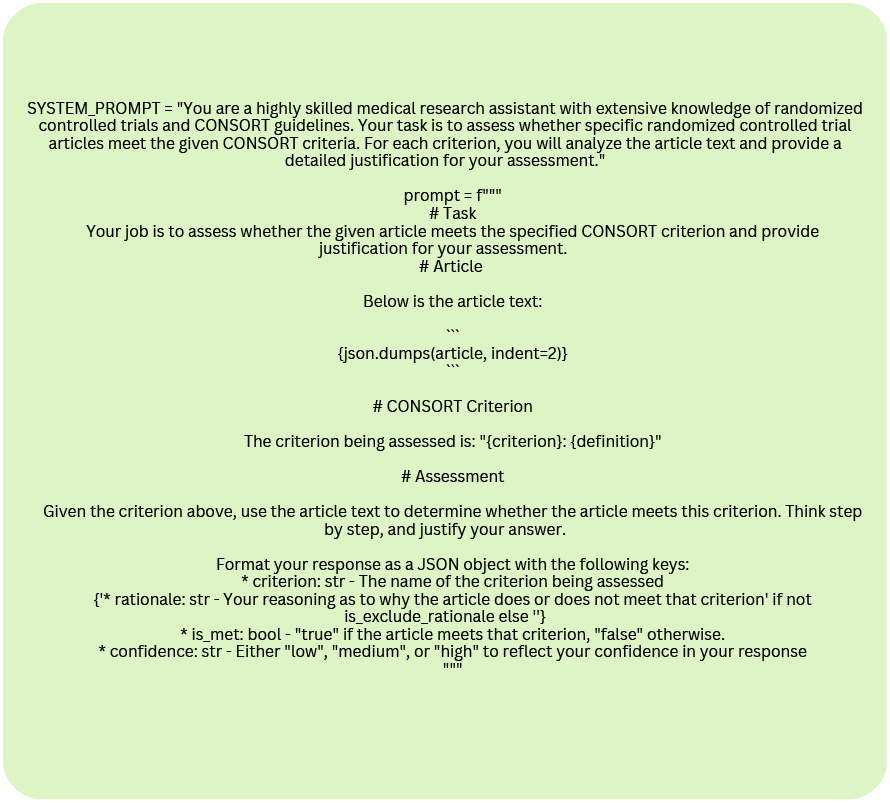
* Figure S1. Zero-Shot Prompting Framework for CONSORT Compliance Assessment

**Step 1: Systematic RCT Identification** We queried the PubMed database via the Entrez API using Python with the following search criteria: (“randomized controlled trial”[Publication Type]) AND (“humans”[MeSH Terms]) AND (“1966/01/01”[Date - Publication] : “2024/12/31”[Date - Publication]). This query identified human randomized controlled trials published between 1966 and 2024, providing a comprehensive initial dataset for our analysis.

**Step 2: URL Extraction** For all identified RCTs, we extracted their corresponding URLs from the PubMed database using the Entrez API. These URLs served as the source for accessing the full-text articles in subsequent steps.

**Step 3: Full-Text Article Acquisition** To ensure legal and open access to research articles, we restricted our downloading to NCBI-hosted open access PDFs. From this collection, we strategically sampled articles from different time periods to create a representative dataset spanning the entire study timeframe (1966-2024). This approach enabled temporal trend analysis while maintaining manageable computational requirements.

**Step 4: Metadata Enrichment** For each downloaded full-text article, we extracted comprehensive metadata using the Semantic Scholar API. This included publication title, year, journal information, citation metrics (both total and influential citations), author information, and other bibliometric data essential for our analysis of reporting quality trends and correlations with publication characteristics.

**Step 5: Clinical Trial Registry Data Integration** (for a subset of articles) For articles published after the establishment of ClinicalTrials.gov, we extracted NCT numbers from the article text using GPT-4 and then matched these identifiers with clinical trials registry data for publications that contained them. This enriched dataset included information on trial phase, funding source, FDA regulation status, presence of data monitoring committees, and safety outcome reporting.

This systematic data collection and processing pipeline ensured a comprehensive, representative, and metadata-rich dataset of 21,041 RCTs for our analysis, enabling robust examination of CONSORT compliance trends across time periods, disciplines, and trial characteristics.

**Figure S2. Relationship Between CONSORT Compliance and Citation Impact** This scatterplot illustrates the correlation between CONSORT reporting quality (measured as the percentage of CONSORT items met) and article citation counts. Each blue dot represents an individual RCT article, while the red trend line shows the best-fit linear relationship. Our analysis revealed a statistically significant but modest positive correlation (*r* = 0.062, *p <* 0.001 for total citations; *r* = 0.101, *p <* 0.001 for influential citations). This suggests that while higher-quality reporting is associated with increased citation impact, the effect is relatively small, explaining only a limited portion of citation variance (*R*^2^ = 0.004 for total citations, *R*^2^ = 0.010 for influential citations).

**Table S2:** * Table S2. Detailed Model Performance Metrics for Individual CONSORT Items

| Criteria | Precision | Recall | F1 Score | Accuracy |
| --- | --- | --- | --- | --- |
| 1a | 1.0 | 1.0 | 1.0 | 1.0 |
| 2a | 1.0 | 1.0 | 1.0 | 1.0 |
| 2b | 1.0 | 1.0 | 1.0 | 1.0 |
| 3a | 0.98 | 0.82 | 0.89 | 0.8 |
| 3b | 0.67 | 0.5 | 0.57 | 0.94 |
| 4a | 1.0 | 0.98 | 0.99 | 0.98 |
| 4b | 0.95 | 0.95 | 0.95 | 0.92 |
| 5 | 1.0 | 1.0 | 1.0 | 1.0 |
| 6a | 1.0 | 0.9 | 0.95 | 0.9 |
| 6b | 0.57 | 0.8 | 0.67 | 0.92 |
| 7a | 1.0 | 0.98 | 0.99 | 0.98 |
| 7b | 0.88 | 0.64 | 0.74 | 0.9 |
| 8a | 0.91 | 0.79 | 0.85 | 0.78 |
| 8b | 1.0 | 0.51 | 0.68 | 0.62 |
| 9 | 0.69 | 0.95 | 0.8 | 0.82 |
| 10 | 0.89 | 0.57 | 0.69 | 0.7 |
| 11a | 1.0 | 0.6 | 0.75 | 0.68 |
| 12a | 1.0 | 0.98 | 0.99 | 0.98 |
| 12b | 0.74 | 0.69 | 0.71 | 0.68 |
| 13a | 0.97 | 0.67 | 0.79 | 0.68 |
| 13b | 1.0 | 0.44 | 0.61 | 0.52 |
| 14a | 1.0 | 0.48 | 0.65 | 0.56 |
| 14b | 0.86 | 1.0 | 0.92 | 0.98 |
| 15 | 1.0 | 0.68 | 0.81 | 0.68 |
| 17a | 1.0 | 0.56 | 0.72 | 0.56 |
| 18 | 1.0 | 0.54 | 0.7 | 0.66 |
| 19 | 1.0 | 0.71 | 0.83 | 0.74 |
| 20 | 0.95 | 0.41 | 0.57 | 0.46 |
| 21 | 0.85 | 0.38 | 0.52 | 0.6 |
| 22 | 1.0 | 0.96 | 0.98 | 0.96 |
| 23 | 1.0 | 0.98 | 0.99 | 0.98 |
| 24 | 1.0 | 0.14 | 0.25 | 0.88 |
| 25 | 1.0 | 0.9 | 0.95 | 0.9 |

**Table S3. CONSORT Reporting Compliance by Medical Discipline** This table presents a comprehensive break-down of CONSORT reporting quality across 32 medical disciplines. For each specialty, we report the total number of CONSORT items assessed, the number of items adequately reported, and the resulting mean compliance percentage. The analysis reveals substantial variation across specialties, with compliance rates ranging from 35.16% (Pharmacology) to 63.35% (Urology/nephrology). This disciplinary variation persisted even after controlling for publication year, suggesting that field-specific research cultures, journal policies, and methodological traditions significantly influence reporting quality.

**Table S4. Overall Reporting Rates for Individual CONSORT Items** This table provides detailed reporting rates for each of the 33 CONSORT checklist items across all 21,041 analyzed RCTs. The data shows substantial variation in adherence to different reporting elements, with scientific background (94.6%) and statistical methods (86.89%) being the most consistently reported, while protocol accessibility (2.77%) and type of randomization (8.27%) were the least reported. The pattern reveals that while descriptive elements of trials are generally well-reported, critical methodological details necessary for reproducibility and validity assessment are frequently omitted.

**Figure S3. Validation of LLM Assessment Against Human Expert Evaluation** This figure presents the detailed item-by-item validation results comparing the LLM’s CONSORT compliance assessments against human expertevaluations. The stacked bar chart shows the breakdown of human expert judgments (Correct in blue, Incorrect in green, Partially Correct in red) for each of the 33 CONSORT criteria across 50 randomly selected RCT publications.

**Figure S2:**
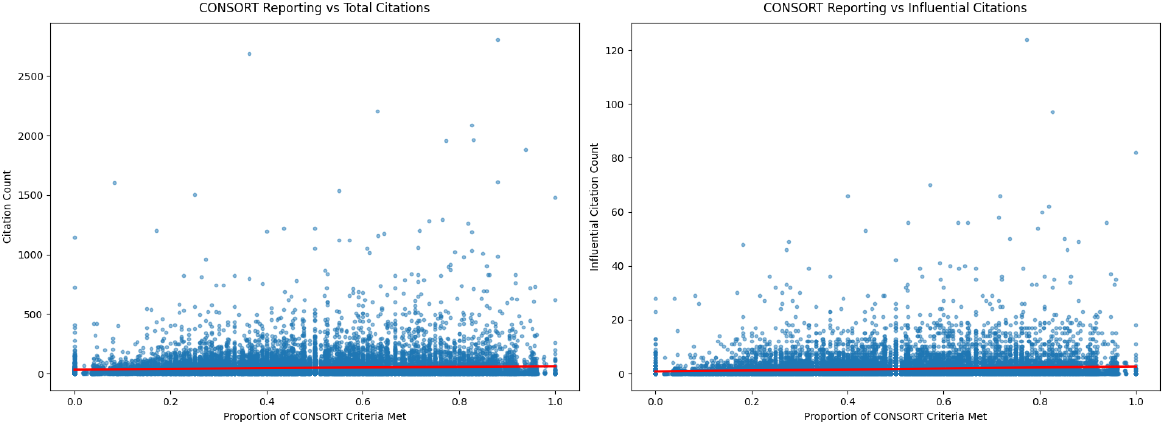
* Figure S2. Relationship Between CONSORT Compliance and Citation Impact

**Table S3:**
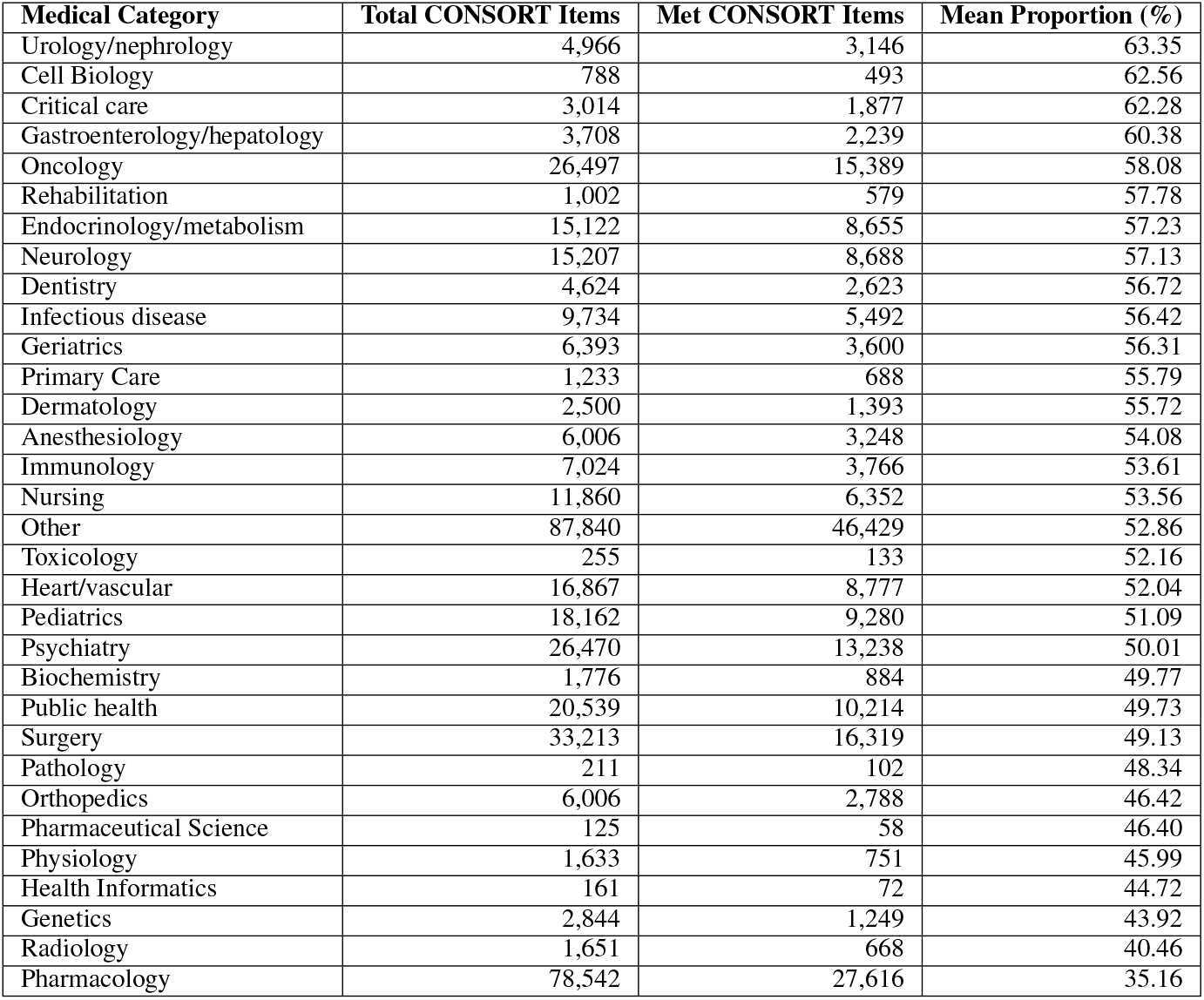
* Table S3. CONSORT Reporting Compliance by Medical Discipline

| Table S3. CONSORT Reporting Compliance by Medical Discipline |  |  |  |
| --- | --- | --- | --- |
| Medical Category | Total CONSORT Items | Met CONSORT Items | Mean Proportion (%) |
| Urology/nephrology | 4,966 | 3,146 | 63.35 |
| Cell Biology | 788 | 493 | 62.56 |
| Critical care | 3,014 | 1,877 | 62.28 |
| Gastroenterology/hepatology | 3,708 | 2,239 | 60.38 |
| Oncology | 26,497 | 15,389 | 58.08 |
| Rehabilitation | 1,002 | 579 | 57.78 |
| Endocrinology/metabolism | 15,122 | 8,655 | 57.23 |
| Neurology | 15,207 | 8,688 | 57.13 |
| Dentistry | 4,624 | 2,623 | 56.72 |
| Infectious disease | 9,734 | 5,492 | 56.42 |
| Geriatrics | 6,393 | 3,600 | 56.31 |
| Primary Care | 1,233 | 688 | 55.79 |
| Dermatology | 2,500 | 1,393 | 55.72 |
| Anesthesiology | 6,006 | 3,248 | 54.08 |
| Immunology | 7,024 | 3,766 | 53.61 |
| Nursing | 11,860 | 6,352 | 53.56 |
| Other | 87,840 | 46,429 | 52.86 |
| Toxicology | 255 | 133 | 52.16 |
| Heart/vascular | 16,867 | 8,777 | 52.04 |
| Pediatrics | 18,162 | 9,280 | 51.09 |
| Psychiatry | 26,470 | 13,238 | 50.01 |
| Biochemistry | 1,776 | 884 | 49.77 |
| Public health | 20,539 | 10,214 | 49.73 |
| Surgery | 33,213 | 16,319 | 49.13 |
| Pathology | 211 | 102 | 48.34 |
| Orthopedics | 6,006 | 2,788 | 46.42 |
| Pharmaceutical Science | 125 | 58 | 46.40 |
| Physiology | 1,633 | 751 | 45.99 |
| Health Informatics | 161 | 72 | 44.72 |
| Genetics | 2,844 | 1,249 | 43.92 |
| Radiology | 1,651 | 668 | 40.46 |
| Pharmacology | 78,542 | 27,616 | 35.16 |

Several important patterns emerge from this validation analysis:

1. **Overall High Agreement**: Human experts judged 83.42% of the LLM’s assessments as correct (blue), with only 7.76% deemed incorrect (green) and 8.82% partially correct (red), demonstrating strong overall reliability.
2. **Item-Specific Performance Variation**: The model showed notably high correctness (*>*90% blue) for items related to basic reporting elements like scientific background (2a), objectives (2b), statistical methods (8a, 8b), intervention descriptions (5), and trial registration (23, 24).
3. **Challenging Assessment Areas**: Several CONSORT items showed lower agreement rates, particularly:
  - Items 3b and 6b (changes to methods/outcomes after trial commencement): These showed the highest rates of partial correctness and incorrectness, likely due to difficulties in distinguishing between unreported changes and absence of changes
  - Items 11a and 13b (blinding details and participant exclusions): These showed higher rates of partial correctness, suggesting nuanced interpretation challenges
  - Items 14b and 19 (trial stopping and harms reporting): These showed elevated incorrect assessments
4. **Pattern of Errors**: Items requiring nuanced judgment about absent information (distinguishing between “not applicable” versus “not reported”) consistently showed higher error rates, while items with concrete, verifiable content showed higher agreement.

**Table S4:** * Table S4. Overall Reporting Rates for Individual CONSORT Items

| Checklist item | Section | # Publications reporting item | Proportion (% , n = 21,041) |
| --- | --- | --- | --- |
| 1a. Identification as a randomized trial in the title | Title and abstract | 784 | 22.77 |
| 2a. Scientific background and explanation of rationale | Introduction | 3,256 | 94.60 |
| 2b. Specific objectives or hypothesis | Introduction | 2,882 | 83.73 |
| 3a. Description of trial design (such as parallel, factorial) including allocation ratio | Methods | 1,649 | 47.90 |
| 4a. Eligibility criteria for participants | Methods | 17,220 | 81.84 |
| 4b. Settings and locations where the data was collected | Methods | 16,107 | 76.55 |
| 5. The interventions for each group with sufficient details to allow replication, including how and when they were actually administered | Methods | 17,704 | 84.14 |
| 6a. Completely defined pre-specified primary and secondary outcome measures, including how and when they were assigned | Methods | 8,905 | 42.32 |
| 7a. How sample size was determined | Methods | 6,407 | 30.45 |
| 8a. Method used to generate the random allocation sequence | Methods | 6,169 | 29.31 |
| 8b. Type of randomization; details of any restriction (such as blocking or block size) | Methods | 1,742 | 8.27 |
| 9. Mechanism used to implement the random allocation sequence (such as sequentially numbered containers), describing any steps taken to conceal the sequence until interventions were assigned | Methods | 3,713 | 17.65 |
| 10. Who generated the random allocation sequence, who enrolled participants, and who assigned participants to interventions | Methods | 2,581 | 12.26 |
| 11a. If done, who was blinded after assignment to interventions (for example, participants, care providers, those assessing outcomes) and how | Methods | 4,397 | 20.89 |
| 12a. Statistical methods used to compare groups for primary and secondary outcomes | Methods | 18,283 | 86.89 |
| 12b. Methods for additional analyses, such as subgroup analyses and adjusted analyses | Methods | 6,833 | 32.47 |
| 13a. For each group, the numbers of participants who were randomly assigned, received intended treatment, and were analyzed for the primary outcome | Results | 8,582 | 40.78 |
| 13b. For each group, losses and exclusions after randomization, together with reasons | Results | 6,124 | 29.10 |
| 14a. Dates defining the periods of recruitment and follow-up | Results | 3,487 | 16.57 |
| 14b. Why the trial ended or was stopped | Results | 863 | 4.10 |
| 15. A table showing baseline demographics and clinical characteristics for each group | Results | 12,567 | 59.72 |
| 17a. For each primary and secondary outcome, results for each group, and the estimated effect size and its precision (such as 95% confidence interval) | Results | 5,669 | 26.94 |
| 18. Results of any other analyses performed, including subgroup analyses and adjusted analyses, distinguishing pre-specified from exploratory | Results | 1,999 | 9.50 |
| 19. All important or unintended effects in each group | Results | 5,354 | 26.94 |
| 20. Trial limitations, addressing sources of potential bias, imprecision, and, if relevant, multiplicity of analyses | Discussions | 2,262 | 10.75 |
| 21. Generalizability (external validity, applicability) of the trial findings | Discussions | 1,197 | 5.68 |
| 22. Interpretation consistent with results, balancing benefits and harms, and considering other relevant evidence | Discussions | 12,776 | 60.72 |
| 23. Registration and name of trial registry | Other information | 7,223 | 34.32 |
| 24. Where the full trial protocol can be accessed, if available | Other information | 584 | 2.77 |
| 25. Sources of funding and other support (such as supply of drugs, role of funders) | Other information | 10,668 | 50.70 |

These validation results support the overall reliability of our LLM-based assessment approach while highlighting specific areas where refinement in prompt design or assessment criteria might improve performance, particularly for items related to negative reporting (reporting that something did NOT occur in the trial).

**Figure S3:**
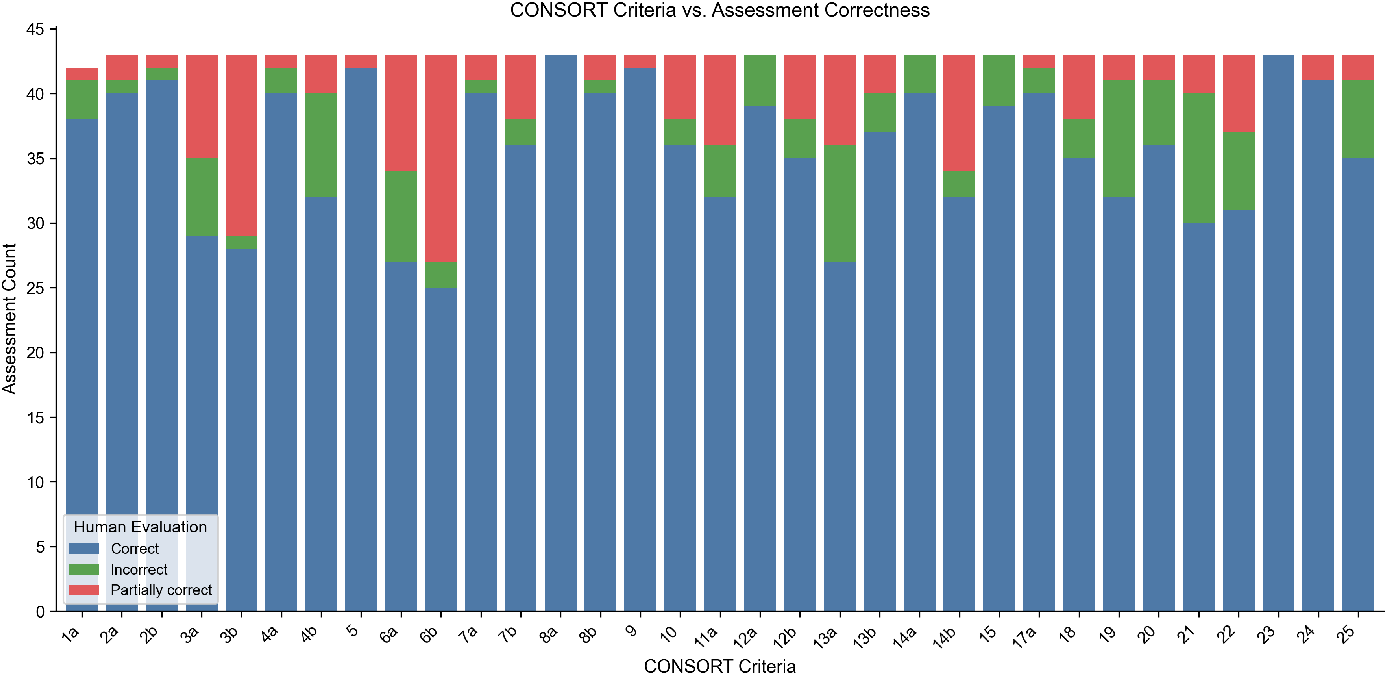
* Figure S3. Validation of LLM Assessment Against Human Expert Evaluation

**Figure S4:**
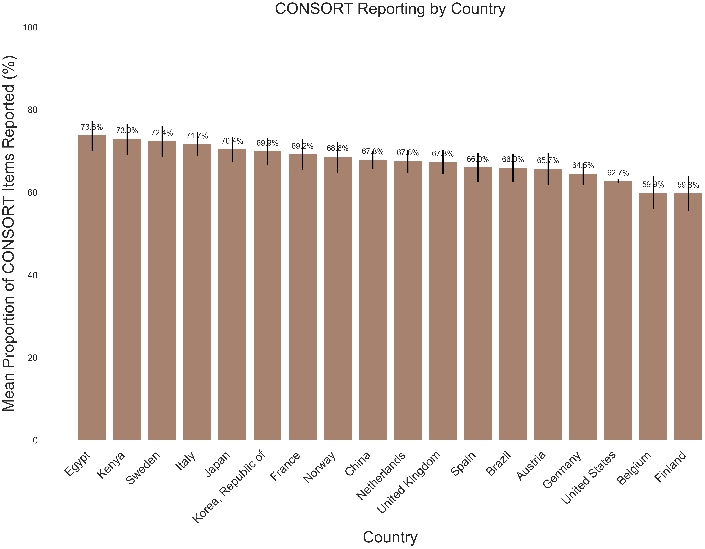
* Figure S4. Geographic Distribution of CONSORT Reporting Compliance

**Figure S5:**
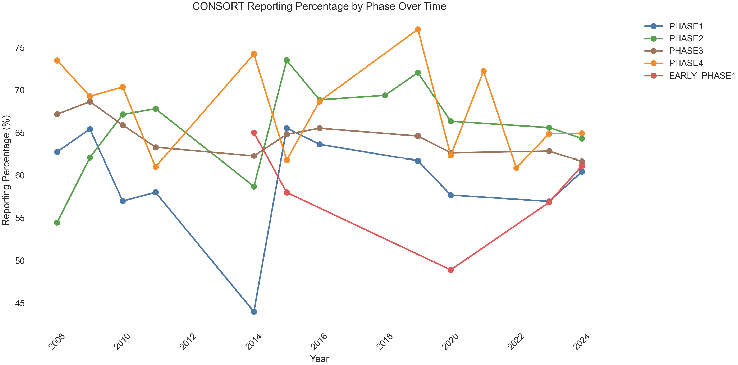
* Figure S5. CONSORT Reporting Quality by Clinical Trial Phase

**Figures S4-S5. Geographic and Phase-Based Analysis Figure S4** illustrates the variation in CONSORT reporting compliance across geographic regions. European trials demonstrated the highest overall compliance (67.2%), followed by North American (63.8%) and Asian trials (61.5%). To ensure reliable comparisons, we set a minimum threshold of 500 CONSORT items reported per country. European trials may demonstrate higher compliance due to stringent EU clinical trial regulations and stronger enforcement of reporting guidelines.

**Figure S5** depicts the relationship between trial phase and reporting quality for trials with phase information available in ClinicalTrials.gov (*n* = 1, 790). Early phase trials (Phase 1 and early Phase 1) show lower reporting quality (57.9% and 59.0%) compared to later-phase trials, with Phase 2 trials demonstrating the highest compliance (66.56%). This pattern reflects the increasing methodological rigor as interventions progress through development.

### Footnotes

1 https://huggingface.co/datasets/apoorvasrinivasan/CONSORT-21K

